# Prevalence, Characteristics and Evolution of Mpox-Related Ophthalmic Disease: A Prospective Cohort Study in South-Kivu, Democratic Republic of the Congo (MBOTE-EYE)

**DOI:** 10.64898/2026.01.29.26344844

**Authors:** Theophile Kabesha, Christophe Van Dijck, Samuel Mudwanga, Sarah Houben, Yves Mujula, Papy Munganga, Jean-Claude Tshomba, Guy Mukari, Tony Wawina-Bokalanga, Eddy Kinganda-Lusamaki, Bart K Jacobs, Achilleas Tsoumanis, Emmanuel Hasivirwe Vakaniaki, Anne W Rimoin, Isabel Brosius, Elise De Vos, Eugene Bangwen, Stefanie Bracke, Jean-Claude Mwanza, Deogratias B Ngoma, Patrick Katoto, Steven Yeh, Nelly N Kabedi, Veronique Nussenblatt, Olivier Tshiani-Mbaya, Ian Crozier, Sabin Sabiti Nundu, Jason Kindrachuk, Laurens Liesenborghs, Placide Mbala-Kingebeni

## Abstract

**Background:** Mpox-related ophthalmic disease (MPOXROD) ranges from mild conjunctivitis to sight-threatening keratitis, however, data based on systematic ophthalmological assessment are scarce. We aimed to characterise the prevalence, features, and temporal evolution of MPOXROD during a clade Ib mpox outbreak.

**Methods:** We conducted MBOTE-EYE, a prospective ophthalmological sub-study, nested within a clinical characterisation cohort in Kamituga, South-Kivu, Democratic Republic of the Congo. All hospitalised patients with mpox confirmed by PCR in the prior 48 hours were eligible. Participants underwent comprehensive ophthalmological examination at enrolment, discharge, and days 29 and 59 post-diagnosis. Conjunctival swabs were collected for monkeypox virus PCR testing. MPOXROD was defined as conjunctivitis, scleritis, keratitis, uveitis, or optic nerve involvement. Risk factors were assessed using mixed-effects Poisson regression.

**Findings:** Between 28 October 2024 and 30 June 2025, 310 participants were enrolled (median age 14 years, IQR 2.5-25.0; 53.5% female, n=166/310). At enrolment, conjunctivitis was present in 36.1% (95% CI 31.0–41.6%, n=112/310), keratitis in 7.7% (95% CI 5.3–11.3%), and anterior uveitis in 0.6% (95% CI 0.2–2.3%). Overall, 43.2% (95% CI 37.8–48.8%) developed MPOXROD during follow-up, most often bilaterally. Visual acuity <8/10 occurred in 22.9% (95% CI 17.6–29.2%, n=24/310), and persistent blindness in 0.9% (95% CI 0.24–5.5%, n=2/225), due to ulcerative keratitis. Periorbital lesions (adjusted risk ratio [aRR] 2.82, 95% CI 1.40–5.69) and severe malnutrition (aRR 5.06, 2.25–11.38) were independently associated with MPOXROD. Conjunctival swabs with PCR Ct values < 25 occurred exclusively in participants with active MPOXROD.

**Interpretation:** Ophthalmic involvement in clade Ib mpox is common and frequently bilateral, with a substantial burden of keratitis and risk of vision loss, particularly in young children and severely malnourished individuals. These findings highlight the need for systematic eye examinations in mpox care and provide critical evidence to inform future trials of targeted ophthalmic therapies.

**Funding:** None of the funders had a role in study design, analysis, interpretation, or writing.

## Introduction

Mpox, caused by monkeypox virus (MPXV) infection, is characterised by fever, skin rash, and lymphadenopathy.^1,2^ The clinical manifestations of mpox can resemble milder forms of smallpox, which was caused by the now eradicated variola virus, another member of the Orthopoxvirus genus that also includes MPXV.^1,2^ Until the last decade, clade I and II MPXV were considered as endemic zoonotic viruses in Central and West Africa, respectively, causing relatively small outbreaks with limited secondary human-to-human transmission.^1,2^ However, since 2017, several large outbreaks of mpox have occurred with extensive human-to-human transmission, mainly driven by sexual contact, and with international spread.^1,3,4^ This has included the declaration of two mpox-related Public Health Emergency of International Concern by the World Health Organization.^5,6^ MPXV clades Ia, Ib, and IIb continue to circulate among dense populations, including sexual networks with over 50,000 cases in 2025, alone.^7^

Although mpox is associated with generally low case fatality and resolution of clinical symptoms and signs within two to three weeks, complications such as bacterial superinfection, large necrotizing skin lesions, dehydration, or proctitis can cause severe disability or death in a small minority of cases, with a higher risk in immunocompromised individuals.^2,8^ Mpox has also been associated with poor pregnancy outcomes.^9^ In addition, mpox-related ophthalmic disease (MPOXROD) has also been described to cause significant morbidity. Studies have reported that MPOXROD can affect 5 to 23% of individuals, ranging from mild conjunctivitis to severe corneal ulceration and visual impairment due to corneal scarring or phthisis bulbi.^10,11^ In the absence of approved antiviral treatments, management is limited to supportive care that includes topical lubrication, anti-inflammatory or antibiotic therapies.^11^

To date, systematic longitudinal characterisation of clade I MPXV-associated ophthalmic manifestations based on expert ophthalmological evaluation has not occurred. Therefore, we aimed to prospectively characterise MPOXROD prevalence, clinical and virologic features and temporal evolution in a cohort of patients infected with clade I MPXV in South-Kivu, the Democratic Republic of the Congo (DRC).

## Materials and methods

### Study design and participants

This study was implemented in Kamituga, South-Kivu (DRC), during an ongoing clade Ib mpox epidemic. All patients with PCR-confirmed mpox were hospitalised in a dedicated mpox treatment centre at Kamituga General Hospital. After receiving medical attention, consenting individuals were enrolled in two parallel observational studies. The MBOTE-Kamituga study (NCT06652646) is an ongoing prospective clinical characterisation study initiated on 2 May 2024, with interim results reported previously.^8^ In addition, from 28 October 2024 onwards, patients of any age or gender with confirmed mpox, defined as a positive MPXV-PCR on a sample collected less than 48 hours before enrolment, could be enrolled in the ophthalmological sub-study, MBOTE-EYE (NCT06579885). The primary objective of MBOTE-EYE was to determine the prevalence of MPOXROD at presentation. Secondary objectives were to describe ophthalmic symptoms and signs, visual acuity, and the presence of MPXV DNA in tear fluid at presentation and during follow-up.

MBOTE-Kamituga and MBOTE-EYE were approved by the Ethics Committee of the University of Kinshasa (approval IDs ESP/CE/78/2024 and ESP/CE/083/2024, respectively) and the University Hospital Antwerp (IDs 6383 and 6440, respectively). MBOTE-EYE was additionally approved by the University of Manitoba Research Ethics Board (ID HS26717). Written consent was obtained before all study procedures from participants aged 18 years and older, and from parents or guardians for minors. Children aged 12–17 years received age-appropriate information and signed an assent form. Specific consent was also obtained for photographs for publication. All participants received reimbursement of transportation costs, and free care and nutrition, including topical ophthalmological therapies.

### Procedures

Participants were investigated by an MBOTE-Kamituga study physician, who collected data on sociodemographics, medical history, symptoms and signs, including WHO severity score^12^. Next, they were examined by a dedicated MBOTE-EYE study ophthalmologist, who collected data on ophthalmic symptoms and signs, including evaluation of visual acuity (optotype test, from the age of three, onward, if understanding of the child allowed); pupillary light reflexes and ocular motility; intraocular pressure (TonoPen® tonometer); portable slit lamp examination of the anterior segment before and after application of fluorescein; and dilated indirect fundoscopic examination and photographic imaging of the eyes and orbits using iPhone 13.

All data were prospectively captured using standardized case report forms in REDCap (Vanderbilt University, Nashville, TN, USA). Between December 19, 2024 and January 10, 2025, MBOTE-Kamituga was paused while MBOTE-EYE continued, resulting in missing data on the medical histories from 48 participants.

The study ophthalmologist also collected conjunctival swabs using minitip FLOQSwabs® of the surface of the lower conjunctival sac. Swabs were immediately placed into 1 mL of universal transport medium and samples were stored at −20°C in small portable field freezers. Regional political instability limited the shipment of materials to and from the study area, necessitating the discontinuation of conjunctival swab collection from December 5, 2024 onward.

Conjunctival swabs were shipped to the Institut National de Recherche Biomédicale, Kinshasa, for PCR analysis using the RADI Fast MPXV Detection Kit (RV015R, KH Medical, Seoul, Republic of Korea), following the manufacturer’s protocol. Cycle threshold (Ct) values were used as semi-quantitative estimates of viral concentrations (higher values corresponding to lower concentrations). Ct values of 40 and above were considered PCR-negative.

Patients were discharged at the discretion of the treating physician. Participants were invited for outpatient follow-up visits at days 29 and 59 post-diagnosis. In case with new or worsening ocular symptoms (reduced visual acuity, photophobia, eye pain, eyelid lesion or oedema) between enrolment and day 59, an additional study visit was performed.

### Statistical analyses

Demographic, clinical, virological and ophthalmological characteristics were summarized using medians with interquartile ranges (IQR) for continuous variables and numerator/denominator (%) for categorical variables. Baseline characteristics were presented by age group reflecting physiological differences: children <5 years of age, children from 5-14 years and individuals >15 years.

Descriptive analyses were conducted at the participant level, classifying ophthalmic conditions as present if observed in either eye. Regression analyses were performed at the eye level, with one observation per eye. MPOXROD was defined as the presence of conjunctivitis, scleritis, keratitis, uveitis or optic nerve involvement. The association between MPOXROD and covariates was determined using a generalized linear mixed-effects model with a Poisson distribution (GLM-P), specifying MPOXROD as a binary outcome and including eye (left/right) nested within participant ID as random intercepts to account for within-subject correlation. We examined the following covariates: age group; sex; presence of periorbital, respectively palpebral mpox lesions; WHO severity score: (total body lesion count <25 (“mild”), 25-99 (“moderate”), 100-250 (“severe”), or >250 (“grave”)); severe malnutrition; PCR-positivity; and the interaction between PCR-positivity and Ct-value. Variables associated with MPOXROD in univariable analyses with a p-value < 0.20 were included in the multivariable model. Next, we used a similar GLM-P with a binary outcome defined as Snellen visual acuity score < 8/10, and MPOXROD as the predictor. Results of the regression models were presented as risk-ratios (RR) with the corresponding 95% confidence interval (95% CI).

Besides the regression modelling, which relied on a complete-case approach, all other analyses were done on an available-case basis, with denominators representing the number of individuals with available data for that variable. Statistical analyses were performed using R software (version 4.4.2).

## Results

Between 28 October 2024 and 30 June 2025, 320 individuals were screened for eligibility, and 310 (96.9%) were enrolled in MBOTE-EYE (**Figure 1**).

**Figure 1:**
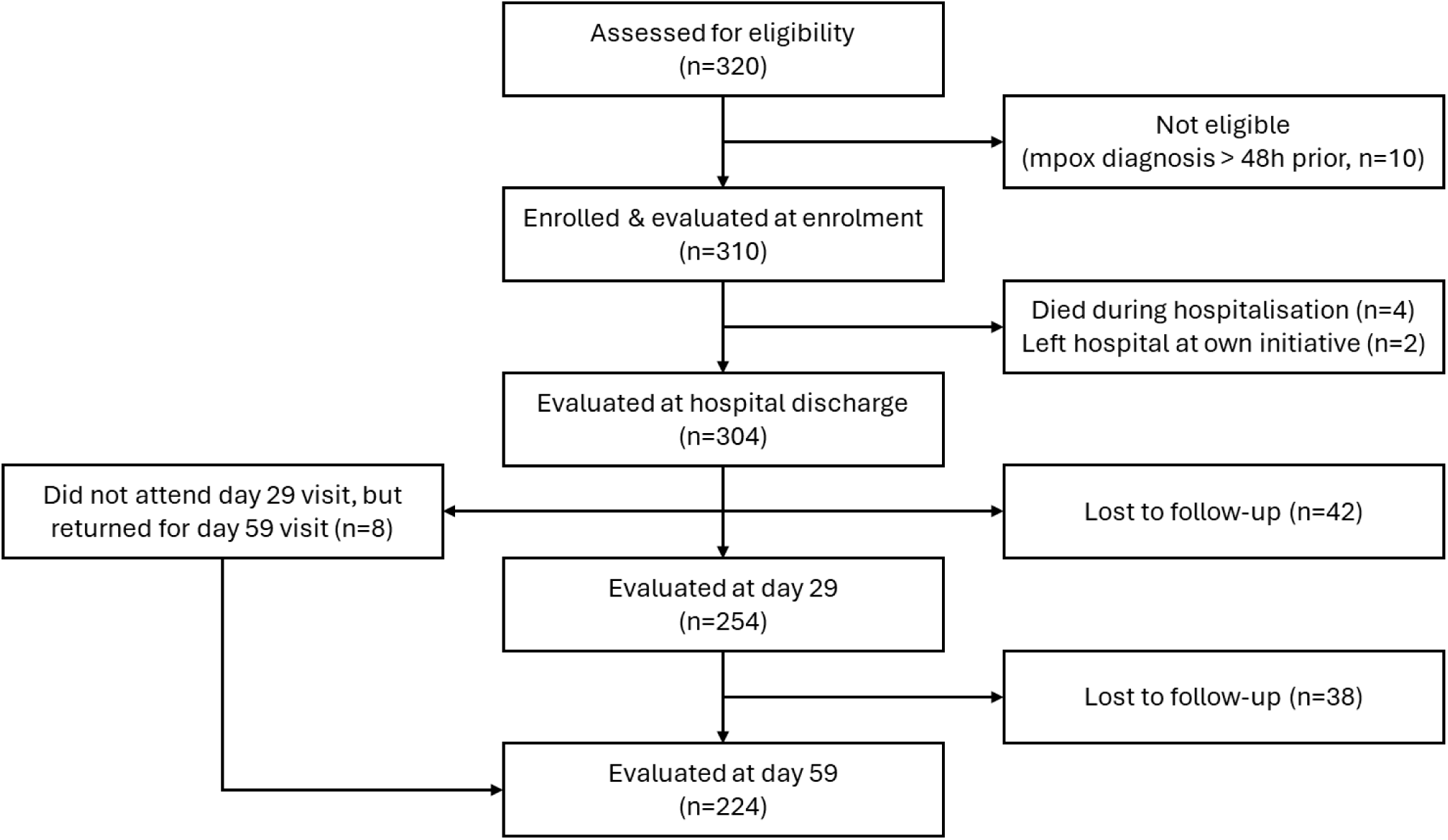
Participant enrolment and follow-up

About half (166/310, 53.5%) were women, and the median age was 14.0 (IQR 2.5-25.0) years. Comorbidities included malnutrition (n=22), HIV infection (n=3), renal disease (n=2), tuberculosis (n=2), hepatic disease (n=1), and diabetes mellitus (n=1, **Table 1**). Twelve participants (12/262, 5.6%) received a vaccinia-based vaccine before the development of mpox symptoms (n=3 smallpox vaccination > 40 years earlier; n=9 Modified Vaccinia-Ankara Bavarian Nordic vaccination in 2025). One in ten participants (37/310, 11.9%) had signs of prior ophthalmic disease at enrolment, including refractive error (n=22), pterygium (n=8), cataract (n=5), glaucoma (n=2), conjunctival melanoma (n=1), and pinguecula (n=1).

**Table 1:** Baseline participant characteristics.

|  | Age < 5<br>(N=105) | Age 5 – 14<br>(N=56) | Age > 14<br>(N=149) | Overall<br>(N=310) |
| --- | --- | --- | --- | --- |
| Median age (IQR) | 1.2 (0.6–2.5) | 8.0 (6.8–12.0) | 25.0 (21.0–30.0) | 14.0 (2.5–25.0) |
| Female sex, n/N (%) | 51/105 (48.6) | 31/56 (55.4) | 84/149 (56.4) | 166/310 (53.5) |
| Missing record in MBOTE-Kamituga study (Christmas period 2024) | 17 | 7 | 24 | 48 |
| Median time (days) since onset of symptoms (IQR) | 6.5 (4.0–8.0) | 5.0 (4.0–7.0) | 7.0 (4.0–9.0) | 6.0 (4.0–8.0) |
| Median time (days) since onset of skin lesions (IQR) | 4.5 (3.0–6.0) | 4.0 (3.0–5.0) | 5.0 (3.0–7.0) | 4.0 (3.0–7.0) |
| **Comorbidities, n/N (%)** |  |  |  |  |
| HIV infection | 0/88 (0.0) | 1/49 (2.0) | 2/125 (1.6) | 3/262 (1.1) |
| Diabetes mellitus | 0/88 (0.0) | 0/49 (0.0) | 1/125 (0.8) | 1/262 (0.4) |
| Tuberculosis | 0/88 (0.0) | 0/49 (0.0) | 2/125 (1.6) | 2/262 (0.8) |
| Hepatic disease | 0/88 (0.0) | 1/49 (2.0) | 0/125 (0.0) | 1/262 (0.4) |
| Renal disease | 0/88 (0.0) | 1/49 (2.0) | 1/125 (0.8) | 2/262 (0.8) |
| Malnutrition<sup>£</sup> |  |  |  |  |
| Moderate | 4/86 (4.7) | 1/49 (2.0) | 10/123 (8.1) | 15/258 (5.8) |
| Severe | 2/86 (2.3) | 3/49 (6.1) | 2/123 (1.6) | 7/258 (2.7) |
| **Vaccination status, n/N (%)** |  |  |  |  |
| Received smallpox vaccination during childhood | 0/88 (0.0) | 0/49 (0.0) | 3/117 (2.6) | 3/254 (1.2) |
| Received MVA-BN vaccination | 0/88 (0.0) | 0/49 (0.0) | 9/125 (7.2) | 9/262 (3.4) |
| **Mpox severity (total number of lesions), n/N (%)** |  |  |  |  |
| Mild (<25 lesions) | 22/88 (25.0) | 11/49 (22.4) | 45/125 (36.0) | 78/262 (29.8) |
| Moderate (25–99 lesions) | 26/88 (29.5) | 15/49 (30.6) | 36/125 (28.8) | 77/262 (29.4) |
| Severe (100–250 lesions) | 19/88 (21.6) | 14/49 (28.6) | 25/125 (20.0) | 58/262 (22.1) |
| Grave (>250 lesions) | 21/88 (23.9) | 9/49 (18.4) | 19/125 (15.2) | 49/262 (18.7) |
| **Prior ophthalmological history in at least one eye, n/N (%)** |  |  |  |  |
| None | 104/105 (99.0) | 54/56 (96.4) | 116/149 (77.9) | 273/310 (88.1) |
| Refractive error | 1/105 (1.0) | 2/56 (3.6) | 19/149 (12.8) | 22/310 (10.6) |
| Pterygium | 0/105 (0.0) | 0/56 (0.0) | 8/149 (5.4) | 8/310 (2.6) |
| Cataract | 0/105 (0.0) | 0/56 (0.0) | 5/149 (3.4) | 5/310 (1.6) |
| Dry eyes | 0/105 (0.0) | 0/56 (0.0) | 4/149 (2.7) | 4/310 (1.3) |
| Glaucoma | 0/105 (0.0) | 0/56 (0.0) | 2/149 (1.3) | 2/310 (0.6) |
| Melanoma | 0/105 (0.0) | 0/56 (0.0) | 1/149 (0.7) | 1/310 (0.3) |
| Pinguecula | 0/105 (0.0) | 0/56 (0.0) | 1/149 (0.7) | 1/310 (0.3) |
| **Used topical ophthalmological therapy < 24h before enrolment, n/N (%)** |  |  |  |  |
| Corticosteroids | 0/105 (0.0) | 1/56 (1.8) | 1/149 (0.7) | 2/310 (0.6) |
| Antibiotics (tetracycline) | 4/105 (3.8) | 0/56 (0.0) | 0/149 (0.0) | 4/310 (1.3) |
| Traditional medicines | 0/105 (0.0) | 0/56 (0.0) | 1/149 (0.7) | 1/310 (0.3) |

IQR = interquartile range; MVA-BN = Modified Vaccinia-Ankara Bavarian Nordic
<sup>£</sup> Moderate malnutrition was defined as MUAC 11.5–12.5 cm in children <5 years; BMI-for-age
z-score –3 to –2 SD in participants aged 5–19 years; and BMI 16–18 kg/m<sup>2</sup> in adults >19 years.
Severe malnutrition was defined as MUAC <11.5 cm in children <5 years; BMI-for-age z-score <-3 SD in participants aged 5–19 years; and BMI <16 kg/m<sup>2</sup> in adults >19 years.

Enrolment occurred at a median of 6.0 (IQR 4.0 – 8.0) days following mpox symptom onset. Mpox severity was mild to moderate (<100 lesions) in 59.2% (155/262) of cases. At enrolment, the most common ophthalmic symptoms were itch (64/310, 20.6%), photophobia (62/310, 20%), redness (39/310, 12.6%), pain (36/310, 11.6%), unsharp vision (24/310, 7.7%), and tearing (17/310, 5.5%, **Table 2**). Other symptoms occurred in less than 5%.

**Table 2:** Presence of ophthalmic signs and symptoms, per participant.

|  | **Inclusion** | **Discharge** | **Day 29** | **Day 59** | **Any time point\*** |
| --- | --- | --- | --- | --- | --- |
| **Number of days post onset of symptoms, median, (IQR)** | 6.0 (4.0–8.0) | 16.0 (13.0–22.0) | 34.0 (32.0–36.0) | 64.0 (62.0–66.0) | NA |
| **Symptoms (n/N, %)** | 154/310 (49.7) | 34/304 (11.2) | 39/254 (15.4) | 22/224 (9.8) | 172/310 (55.5) |
| Unsharp vision | 24/310 (7.7) | 15/304 (4.9) | 9/254 (3.5) | 7/224 (3.1) | 29/310 (9.4) |
| Eye pain | 36/310 (11.6) | 3/304 (1.0) | 5/254 (2.0) | 3/224 (1.3) | 42/310 (13.5) |
| Dry eyes | 3/310 (1.0) | 1/304 (0.3) | 0/254 (0.0) | 0/224 (0.0) | 5/310 (1.6) |
| Tears | 17/310 (5.5) | 4/304 (1.3) | 4/254 (1.6) | 4/224 (1.8) | 26/310 (8.4) |
| Feeling of foreign object | 10/310 (3.2) | 2/304 (0.7) | 3/254 (1.2) | 3/224 (1.3) | 16/310 (5.2) |
| Photophobia | 62/310 (20.0) | 9/304 (3.0) | 11/254 (4.3) | 4/224 (1.8) | 75/310 (24.2) |
| Itch | 64/310 (20.6) | 6/304 (2.0) | 12/254 (4.7) | 10/224 (4.5) | 79/310 (25.5) |
| Vision loss | 0/310 (0.0) | 0/304 (0.0) | 0/254 (0.0) | 2/224 (0.9) | 2/310 (0.6) |
| Redness | 39/310 (12.6) | 7/304 (2.3) | 8/254 (3.1) | 5/224 (2.2) | 46/310 (14.8) |
| Discharge or pseudo membranes | 31/310 (10.0) | 1/304 (0.3) | 0/254 (0.0) | 3/224 (1.3) | 34/310 (11.0) |
| Other | 13/310 (4.2) | 1/304 (0.3) | 2/254 (0.8) | 1/224 (0.4) | 18/310 (5.8) |
| **Abnormal eye motility, n/N (%)** | 0/310 (0.0) | 1/304 (0.3) | 1/254 (0.4) | 0/224 (0.0) | 2/310 (0.6) |
| **Mpox lesions** |  |  |  |  |  |
| Periorbital, n/N (%) | 78/310 (25.2) | 0/304 (0.0) | 0/254 (0.0) | 0/224 (0.0) | 79/310 (25.5) |
| Number (median, IQR) | 4.0 (2.0–9.0) | NA | NA | NA | 4.0 (2.0–9.0) |
| On the palpebra, n/N (%) | 60/310 (19.4) | 1/304 (0.3) | 0/254 (0.0) | 1/224 (0.4) | 66/310 (21.3) |
| Number (median, IQR) | 3.0 (2.0–3.5) | 1.0 (1.0–1.0) | NA | 1.0 (1.0–1.0) | 3.0 (1.0–4.0) |
| On the conjunctiva, n/N (%) | 10/310 (3.2) | 4/304 (1.3) | 3/254 (1.2) | 1/224 (0.4) | 13/310 (4.2) |
| Number (median, IQR) | 2.0 (2.0–2.8) | NA | NA | NA | 2.0 (2.0–2.8) |
| **Palpebral edema, n/n (%)** | 18/310 (5.8) | 4/304 (1.3) | 3/254 (1.2) | 3/224 (1.3) | 23/310 (7.4) |
| **Periorbital edema, n/n (%)** | 0/310 (0.0) | 0/304 (0.0) | 0/254 (0.0) | 0/224 (0.0) | 0/310 (0.0) |
| **Conjunctivitis, n/N (%)** | 112/310 (36.1) | 20/304 (6.6) | 26/254 (10.2) | 14/224 (6.2) | 132/310 (42.6) |
| Mild, n/N (%) | 81/108 (75.0) | 15/16 (93.8) | 18/23 (78.3) | 11/13 (84.6) | 106/127 (83.5) |
| Severe, n/N (%) | 25/108 (23.1) | 0/16 (0.0) | 4/23 (17.4) | 2/13 (15.4) | 29/127 (22.8) |
| Conjunctival mpox lesions, n/N (%) | 10/112 (8.9) | 4/20 (20.0) | 3/26 (11.5) | 1/14 (7.1) | 13/132 (9.8) |
| **Scleritis, n/N (%)** | 0/119 (0.0) | 0/304 (0.0) | 0/254 (0.0) | 0/224 (0.0) | 0/310 (0.0) |
| **Corneal abnormality, n/N (%)** | 24/310 (7.7) | 9/304 (3.0) | 8/254 (3.1) | 4/224 (1.8) | 29/310 (9.4) |
| Corneal opacity or scarification, n/N (%) | 2/24 (8.3) | 0/9 (0.0) | 0/8 (0.0) | 2/4 (50.0) | 4/29 (13.8) |
| Corneal epithelial defect or ulcer, n/N (%) | 22/24 (91.7) | 9/9 (100.0) | 8/8 (100.0) | 2/4 (50.0) | 27/29 (93.1) |
| Central, n/N (%) | 12/22 (54.5) | 3/9 (37.5) | 1/8 (12.5) | 2/2 (100.0) | 13/27 (48.1) |
| Peripheral, n/N (%) | 16/22 (72.7) | 6/9 (75.0) | 7/8 (87.5) | 0/1 (0.0) | 24/27 (88.9) |

|  | Inclusion | Discharge | Day 29 | Day 59 | Any time point* |
| --- | --- | --- | --- | --- | --- |
| Corneal edema, n/N (%) | 0/24 (0.0) | 0/9 (0.0) | 0/8 (0.0) | 1/4 (25.0) | 1/29 (3.4) |
| **Hypopion, n/N (%)** | 1/310 (0.3) | 0/304 (0.0) | 0/254 (0.0) | 1/224 (0.4) | 2/310 (0.6) |
| **Abnormality of the iris, n/N (%)** | 2/310 (0.6) | 2/304 (0.7) | 1/254 (0.4) | 1/224 (0.4) | 2/310 (0.6) |
| Synechiae, n/N (%) | 1/2 (50.0) | 1/2 (50.0) | 1/1 (100.0) | 1/1 (100.0) | 1/2 (50.0) |
| Transillumination defects, n/N (%) | 1/2 (50.0) | 1/2 (50.0) | 0/1 (0.0) | 0/1 (0.0) | 1/2 (50.0) |
| **Abnormality of the pupil, n/N (%)** | 1/310 (0.3) | 1/304 (0.3) | 0/254 (0.0) | 2/224 (0.9) | 3/310 (1.0) |
| Semi-mydriasis or Mydriasis, n/N (%) | 0/1 (0.0) | 0/1 (0.0) | 0/0 (NA) | 1/2 (50.0) | 1/3 (33.3) |
| Sluggish or absent photomotor reflex, n/N (%) | 1/1 (100.0) | 1/1 (100.0) | 0/0 (NA) | 1/2 (50.0) | 2/3 (66.7) |
| **Raised intraocular pressure (Tonopen value > 21 mmHg), n/N (%)** | 1/310 (0.3) | 0/304 (0.0) | 0/254 (0.0) | 0/224 (0.0) | 1/310 (0.3) |
| **Cataract, n/N (%)** | 5/310 (1.6) | 3/304 (1.0) | 3/254 (1.2) | 2/224 (0.9) | 5/310 (1.6) |
| **Any vitreous abnormality, n/N (%)** | 1/310 (0.3) | 0/304 (0.0) | 0/254 (0.0) | 1/224 (0.4) | 2/310 (0.6) |
| Opaque vitreous humor, n/N (%) | 1/1 (100.0) | 0/0 (NA) | 0/0 (NA) | 1/1 (100.0) | 2/2 (100.0) |
| **Any abnormality of the optic nerve, n/N (%)** | 2/310 (0.6) | 2/304 (0.7) | 2/254 (0.8) | 0/224 (0.0) | 2/310 (0.6) |
| Optic nerve pallor, n/N (%) | 2/2 (100.0) | 2/2 (100.0) | 2/2 (100.0) | 0/0 (NA) | 2/2 (100.0) |
| **Any macular abnormality, n/N (%)** | 0/310 (0.0) | 0/304 (0.0) | 0/254 (0.0) | 0/224 (0.0) | 0/310 (0.0) |
| **Any retinal vascular abnormality, n/N (%)** | 0/310 (0.0) | 0/304 (0.0) | 0/254 (0.0) | 0/224 (0.0) | 0/310 (0.0) |
| **Any peripheral retinal abnormality, n/N (%)** | 0/310 (0.0) | 0/304 (0.0) | 0/254 (0.0) | 0/224 (0.0) | 0/310 (0.0) |
| **Visual acuity (far sight, without correction)** |  |  |  |  |  |
| Snellen Chart used, n/N (%) | 215/310 (69.4) | 214/304 (70.4) | 170/254 (66.9) | 143/224 (63.8) | 218/310 (70.3) |
| < 1/20 (blindness), n/N (%) | 2/215 (0.9) | 1/214 (0.5) | 1/170 (0.6) | 3/143 (2.1) | 4/218 (1.8) |
| [1/20 - 1/10] (Mediocre Vision), n/N (%) | 0/215 (0.0) | 0/214 (0.0) | 0/170 (0.0) | 0/143 (0.0) | 2/218 (0.9) |
| [1/10 - 3/10] (Poor Vision), n/N (%) | 1/215 (0.5) | 2/214 (0.9) | 2/170 (1.2) | 3/143 (2.1) | 6/218 (2.8) |
| [3/10 - 8/10] (Acceptable Vision), n/N (%) | 44/215 (20.5) | 38/214 (17.8) | 34/170 (20.0) | 26/143 (18.2) | 47/218 (21.6) |
| [8/10 - 10/10] (Good Vision), n/N (%) | 168/215 (78.1) | 173/214 (80.8) | 133/170 (78.2) | 111/143 (77.6) | 177/218 (81.2) |
| Impossibility to use Snellen Chart, n/N (%) | 95/310 (30.6) | 90/304 (29.6) | 84/254 (33.1) | 81/224 (36.2) | 308/310 (99.4) |
| Good light tracking, n/N (%) | 95/95 (100.0) | 90/90 (100.0) | 83/83 (100.0) | 81/81 (100.0) | 99/99 (100.0) |
| **Visual acuity (far sight, with correction)** |  |  |  |  |  |
| Snellen Chart used, n/N (%) | 8/310 (2.6) | 4/304 (1.3) | 2/254 (0.8) | 0/224 (0.0) | 10/310 (3.2) |
| [3/10 - 8/10] (Acceptable Vision), n/N (%) | 0/8 (0.0) | 1/4 (25.0) | 0/2 (0.0) | 0/0 (NA) | 1/10 (10.0) |
| [8/10 - 10/10] (Good Vision), n/N (%) | 8/8 (100.0) | 3/4 (75.0) | 2/2 (100.0) | 0/0 (NA) | 9/10 (90.0) |
| **Visual acuity (near sight, without correction)** |  |  |  |  |  |
| Parinaud scale used, n/N (%) | 199/310 (64.2) | 204/304 (67.1) | 163/254 (64.2) | 145/224 (64.7) | 223/310 (71.9) |
| P2 | 192/199 (96.5) | 198/204 (97.1) | 156/163 (95.7) | 138/145 (95.2) | 217/223 (97.3) |
| P3 | 1/199 (0.5) | 1/204 (0.5) | 2/163 (1.2) | 2/145 (1.4) | 2/223 (0.9) |
| P4 | 1/199 (0.5) | 1/204 (0.5) | 2/163 (1.2) | 1/145 (0.7) | 2/223 (0.9) |
| P5 | 5/199 (2.5) | 4/204 (2.0) | 3/163 (1.8) | 4/145 (2.8) | 5/223 (2.2) |
| Impossibility to use Parinaud scale, n/N (%) | 111/310 (35.8) | 98/304 (32.2) | 88/254 (34.6) | 79/224 (35.3) | 115/310 (37.1) |
| Good | 109/111 (98.2) | 98/98 (100.0) | 88/88 (100.0) | 79/79 (100.0) | 114/115 (99.1) |
| Poor | 1/111 (0.9) | 0/98 (0.0) | 0/88 (0.0) | 0/79 (0.0) | 1/115 (0.9) |
| Impossible to evaluate | 1/111 (0.9) | 0/98 (0.0) | 0/88 (0.0) | 0/79 (0.0) | 1/115 (0.9) |
| **Visual acuity (near sight, with correction)** |  |  |  |  |  |
| Parinaud scale used, n/N (%) | 7/310 (2.3) | 5/304 (1.6) | 7/254 (2.8) | 7/224 (3.1) | 8/310 (2.6) |
| P2 | 6/7 (85.7) | 5/5 (100.0) | 7/7 (100.0) | 7/7 (100.0) | 8/8 (100.0) |
| P5 | 1/7 (14.3) | 0/5 (0.0) | 0/7 (0.0) | 0/7 (0.0) | 1/8 (12.5) |

\*This includes findings at unplanned visits, not displayed in a separate column

Among participants at enrolment, one fourth (78/310, 25.2%) had periorbital and one fifth (60/310, 19.4%) had palpebral mpox lesions (**Figure 2A-B**). The prevalence of conjunctivitis was 36.1% (112/310, 95% CI 31–41.6%), keratitis 7.7% (24/310, 95% CI 5.3–11.3%), and anterior uveitis 0.6% (2/310, 95% CI 0.2–2.3%, **Table 3, Supplementary Table 1, Figure 2A-B**). Two (2/310, 0.6%, 95% CI 0.11–2.6%) participants had optic nerve pallor associated with pre-existing glaucoma. No other participants had posterior segment abnormalities.

**Figure 2.**
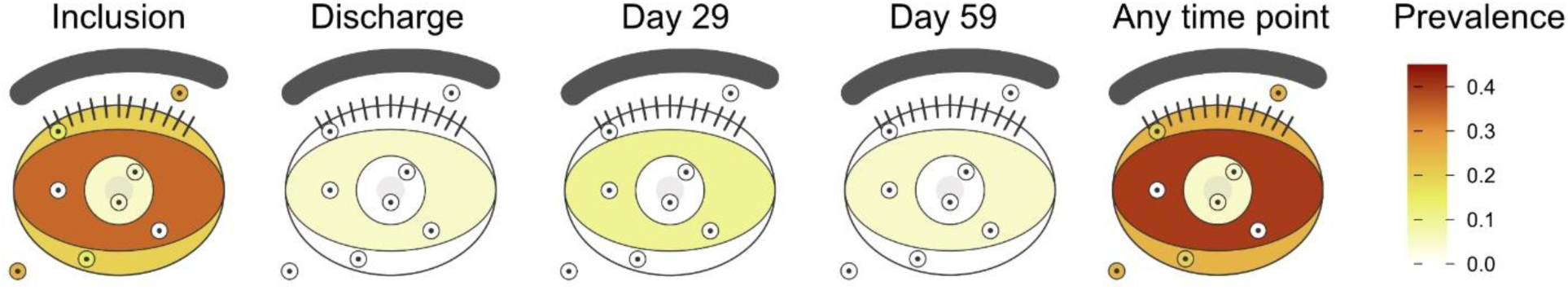
Mpox-related ophthalmic disease: Anatomical heatmap showing prevalence of signs of inflammation (surface fill) or mpox lesions/ulcers (small circles) on the cornea, conjunctiva, palpebra, and periorbita in all study participants over time.

**Table 3:** Ophthalmic outcomes, per participant.

|  | Inclusion | Discharge | Day 29 | Day 59 | Any time point* |
| --- | --- | --- | --- | --- | --- |
| **Median time (days) since onset of symptoms, (IQR)** | 6.0 (4.0–8.0) | 16.0 (13.0–22.0) | 34.0 (32.0–36.0) | 64.0 (62.0–66.0) | NA |
| **Affection of periorbital & palpebra, n/N (%; 95% CI)** |  |  |  |  |  |
| Periorbital mpox lesions | 78/310 (25.2; 20.7–30.3) | 0/304 (0; 0–1.2) | 0/254 (0.0; 0–1.5) | 0/224 (0.0; 0–1.7) | 79/310 (25.5; 21–30.6) |
| Blepharitis | 20/310 (6.5; 4.2–9.8) | 4/304 (1.3; 0.5–3.3) | 3/254 (1.2; 0.4–3.4) | 3/224 (1.3; 0.5–3.9) | 20/310 (6.5; 4.2–9.8) |
| Palpebral mpox lesions | 60/310 (19.4; 15.3–24.1) | 1/304 (0.3; 0–1.8) | 0/254 (0.0; 0–1.5) | 1/224 (0.4; 0–2.5) | 66/310 (21.3; 17.1–26.2) |
| **MPOXROD (mpox-related ocular disease), n/N (%; 95% CI)** | 118/310 (38.1; 32.8–43.6) | 23/304 (7.6; 5.1–11.1) | 29/254 (11.4; 8.1–15.9) | 16/224 (7.1; 4.4–11.3) | 134/310 (43.2; 37.8–48.8) |
| Conjunctivitis | 112/310 (36.1; 31–41.6) | 20/304 (6.6; 4.3–9.9) | 26/254 (10.2; 7.1–14.6) | 14/224 (6.2; 3.8–10.2) | 132/310 (42.6; 37.2–48.1) |
| Without conjunctival mpox lesion(s) | 102/112 (91.1; 84.3–95.1) | 16/20 (80.0; 58.4–91.9) | 23/26 (88.5; 71–96) | 13/14 (92.9; 68.5–99.6) | 119/132 (90.2; 83.9–94.2) |
| With conjunctival mpox lesion(s) | 10/112 (8.9; 4.9–15.7) | 4/20 (20.0; 8.1–41.6) | 3/26 (11.5; 4–29) | 1/14 (7.1; 0.4–31.5) | 13/132 (9.8; 5.8–16.1) |
| Scleritis | 0/310 (0; 0–1.2) | 0/304 (0; 0–1.2) | 0/254 (0; 0–1.5) | 0/224 (0; 0–1.7) | 0/310 (0; 0–1.2) |
| Keratitis | 24/310 (7.7; 5.3–11.3) | 9/304 (3.0; 1.6–5.5) | 8/254 (3.2; 1.6–6.1) | 4/224 (1.8; 0.7–4.5) | 29/310 (9.4; 6.6–13.1) |
| Non ulcerative | 2/24 (8.3; 2.3–25.8) | 0/9 (0; 0–29.9) | 0/8 (0; 0–32.4) | 2/4 (50.0; 15.0–85.0) | 2/29 (6.9; 1.9–22.0) |
| Ulcerative | 22/24 (91.7; 74.2–97.7) | 9/9 (100.0; 70.1–100.0) | 8/8 (100.0; 67.6–100.0) | 2/4 (50.0; 15.0–85.0) | 27/29 (93.1; 78–98.1) |
| Anterior uveitis | 2/310 (0.6; 0.2–2.3) | 2/304 (0.7; 0.2–2.4) | 1/254 (0.4; 0–2.2) | 1/224 (0.4; 0–2.5) | 2/310 (0.6; 0.2–2.3) |
| Intermediate/posterior uveitis | 0/310 (0; 0–1.2) | 0/304 (0; 0–1.2) | 0/254 (0; 0–1.5) | 0/224 (0; 0–1.7) | 0/310 (0; 0–1.2) |
| Affection of the optic nerve | 0/308 (0; 0–1.2) <sup>§</sup> | 0/302 (0; 0–1.3) <sup>§</sup> | 0/252 (0; 0–1.5) <sup>§</sup> | 0/222 (0; 0–1.7) <sup>§</sup> | 0/308 (0; 0–1.2) <sup>§</sup> |
| **Functional outcome, n/N (%; 95% CI)** |  |  |  |  |  |
| Snellen score < 8/10 | 47/215 (21.9; 16.6–28.1) | 41/214 (19.2; 14.2–25.2) | 37/170 (21.8; 16.0–28.9) | 32/143 (22.4; 16.0–30.3) | 50/218 (22.9; 17.6–29.2) |
| Snellen score < 3/10 | 3/215 (1.4; 0.36–4.4) | 3/214 (1.4; 0.36–4.4) | 3/170 (1.8; 0.46–5.5) | 6/143 (4.2; 1.7–9.3) | 7/218 (3.2; 1.4–6.8) |

\*This includes findings at unplanned visits, not displayed in a separate column
<sup>§</sup> Excluding two participants with a prior history of glaucoma and pallor of the optic nerve at enrolment

During the study, participants received topical treatments, including antibiotics (96/310, 31.0%), sodium cromoglycate (57/310, 18.4%), nonsteroidal anti-inflammatory drugs (52/310, 16.8%), artificial tears (28/310, 9.0%), and prednisolone (24/310, 7.7%, **Table 4**). No antivirals were available. Hospital admission had a median duration of 11 (IQR 8.0 – 15.8) days. Ocular signs and symptoms usually improved during hospitalisation, except among 18 participants who were reevaluated because of new or increasing ocular pain (n=10), palpebral oedema or mpox lesions (n=9), reduced visual acuity (n=4), or photophobia (n=2). During such unscheduled visits, eight additional patients were diagnosed with conjunctivitis, five with palpebral mpox lesions, and two with keratitis. Four persons (all < 5 years old) died prior to hospital discharge and two left the hospital at their own initiative. The remaining 304 (98.1%) were examined at hospital discharge, at which time conjunctivitis had resolved in 100/117 (85.5%) of individuals with conjunctivitis, keratitis in 16/25 (64.0%) of those with keratitis, and anterior uveitis persisted in the two affected individuals.

**Table 4:** Topical treatments prescribed throughout the study.

| Medicine, n/N (%) | Overall (N = 310) | Keratitis (N = 29) | Conjunctivitis (N = 132) | Anterior uveitis (N = 2) |
| --- | --- | --- | --- | --- |
| Antibiotic eye drops | 88/310 (28.4) | 27/29 (93.1) | 75/132 (56.8) | 0/2 (0.0) |
| Sodium cromoglycate eye drops | 57/310 (18.4) | 5/29 (17.2) | 55/132 (41.7) | 0/2 (0.0) |
| NSAID eye drops | 52/310 (16.8) | 23/29 (79.3) | 42/132 (31.8) | 0/2 (0.0) |
| Tetracycline ointment | 43/310 (13.9) | 21/29 (72.4) | 33/132 (25.0) | 0/2 (0.0) |
| Artificial tears | 28/310 (9.0) | 4/29 (13.8) | 16/132 (12.1) | 0/2 (0.0) |
| Prednisolone eye drops | 24/310 (7.7) | 4/29 (13.8) | 22/132 (16.7) | 2/2 (100.0) |
| Acyclovir eye drops | 2/310 (0.6) | 2/29 (6.9) | 2/132 (1.5) | 0/2 (0.0) |
| Atropine eye drops | 2/310 (0.6) | 1/29 (3.4) | 1/132 (0.8) | 2/2 (100.0) |
| Dexamethasone-gentamycin injection (sub-conjunctival) | 2/310 (0.6) | 1/29 (3.4) | 1/132 (0.8) | 1/2 (50.0) |
| Beta antagonist eye drops | 2/310 (0.6) | 0/29 (0.0) | 2/132 (1.5) | 0/2 (0.0) |

Among participants attending outpatient follow-up, conjunctivitis was observed in 10.2% (26/254, 95% CI 7.1–14.6%), and keratitis in 3.2% (8/254, 95% CI 0.7–4.5%) at day 29, decreasing to 6.2% (14/224, 95% CI 3.8–10.2%) and 1.8% (4/224, 95% CI 0.7–4.5%), respectively, at day 59. Anterior uveitis persisted in one participant at both timepoints (1/254, 0.4%, 95% CI 0–2.2% at day 29 and 1/224, 0.4%, 95% CI 0–2.5% at day 59). The second study participant diagnosed with anterior uveitis was lost to follow-up after hospital discharge.

Overall, during the observation period, 43.2% (134/310, 95% CI 37.8–48.8%) developed MPOXROD, in whom 3.7% (5/134) and 96.3% (129/134) had unilateral and bilateral disease, respectively. Signs of MPOXROD were often already present at enrolment (215/263, 81.7% of eyes). Within the limits of our observation period, although keratitis frequently co-occurred with conjunctivitis at some stage (37/47, 78.7% of eyes), it was rarely preceded by a phase of milder eye disease such as conjunctivitis (8/47, 17.0% of eyes). Among the participants for whom Snellen charts could be used, visual acuity scored below 8/10 at least once during follow-up in 22.9% (50/218, 95% CI 17.6–29.2%) and below 3/10 in 3.2% (7/218, 95% CI 1.4–6.8%). Two individuals (2/225, 0.9%, 95% CI 0.24–5.5%) had persistent blindness (Snellen score < 1/20) at day 59 resulting from ulcerative keratitis (**Supplementary Figure 1**). Active MPOXROD was a significant risk factor for a visual acuity score < 8/10 (RR 1.20, 95% CI 1.03–1.41, **Figure 3**).

**Figure 3:**
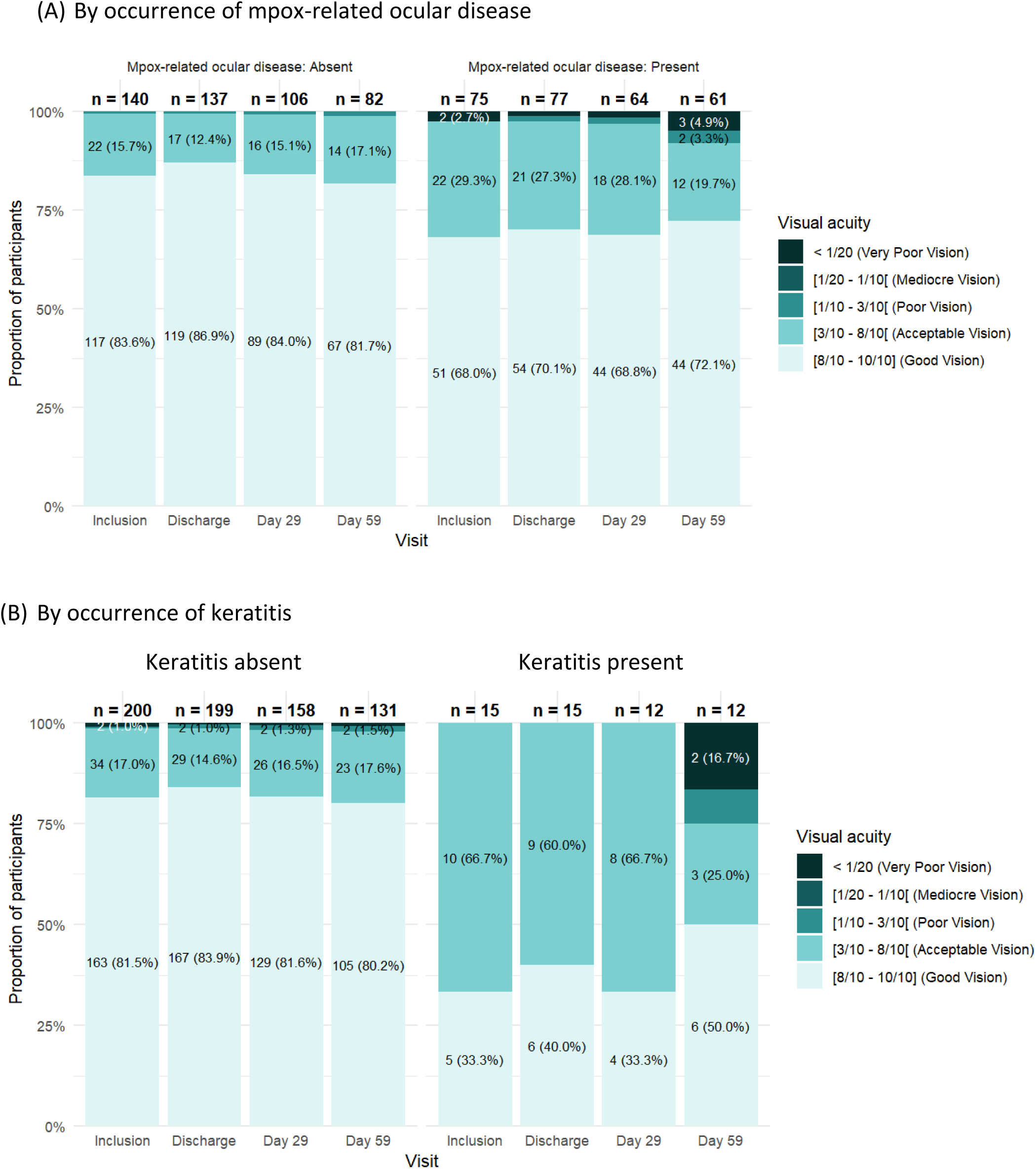
Visual acuity score over time among individuals who could be evaluated using a Snellen Chart

MPXV-PCR results were available from 312 conjunctival swabs of 89 participants: 132/178 (74.2%) swabs from 89 participants were PCR-positive at enrolment, with median PCR Ct values of 31.7 (IQR 28.9–34.7), 65/104 (62.5) were PCR-positive at discharge with median Ct values of 33.8 (IQR 32.7–35.3), and 9/30 (30.0%) were PCR-positive at day 29 with median Ct values of 35.0 (IQR 34.7–36.4, **Figure 4A-B, Supplementary Table 2**). No swabs were collected at day 59. PCR Ct values below 25 were observed exclusively among individuals with active signs of MPOXROD.

**Figure 4:**
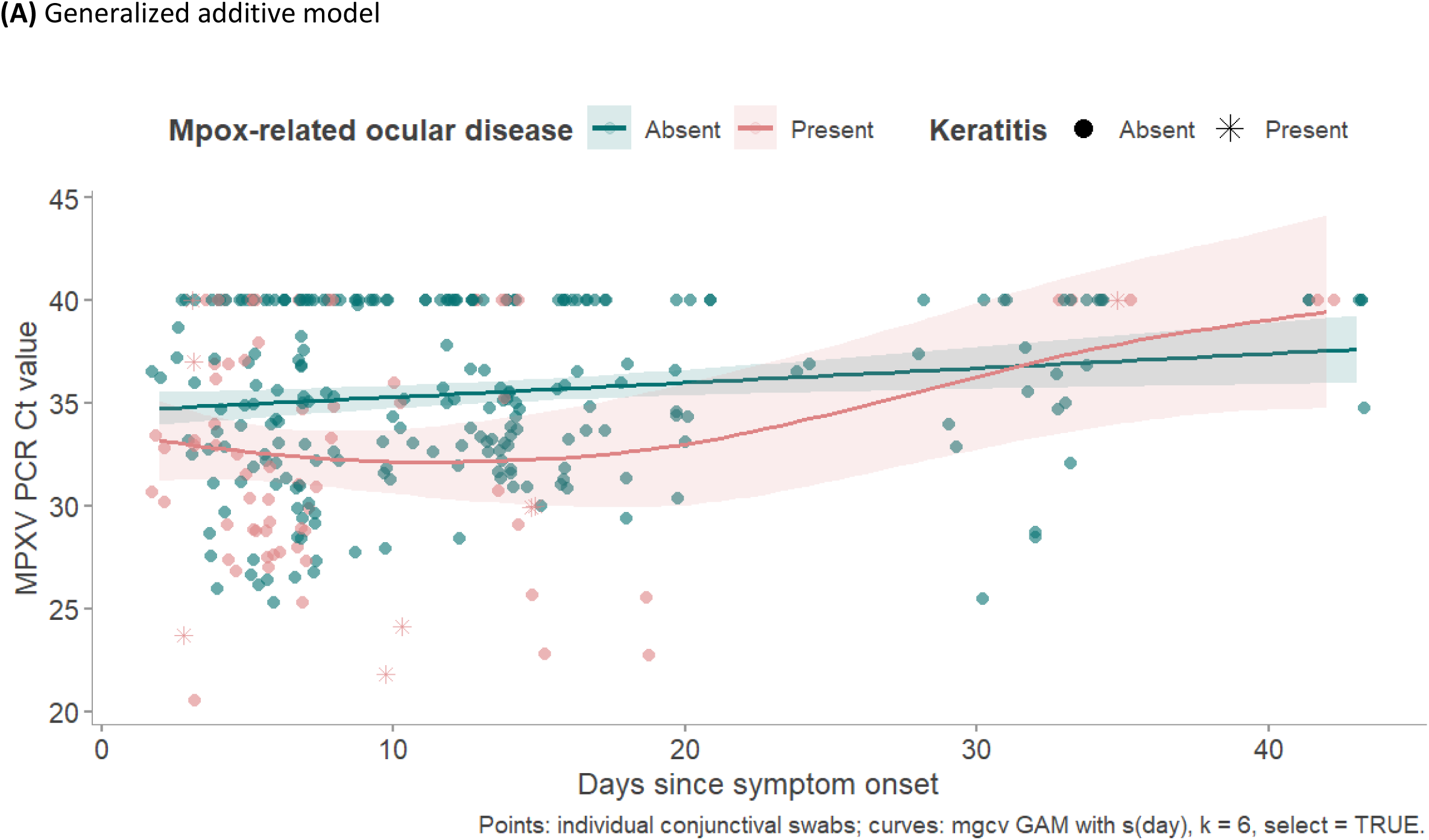
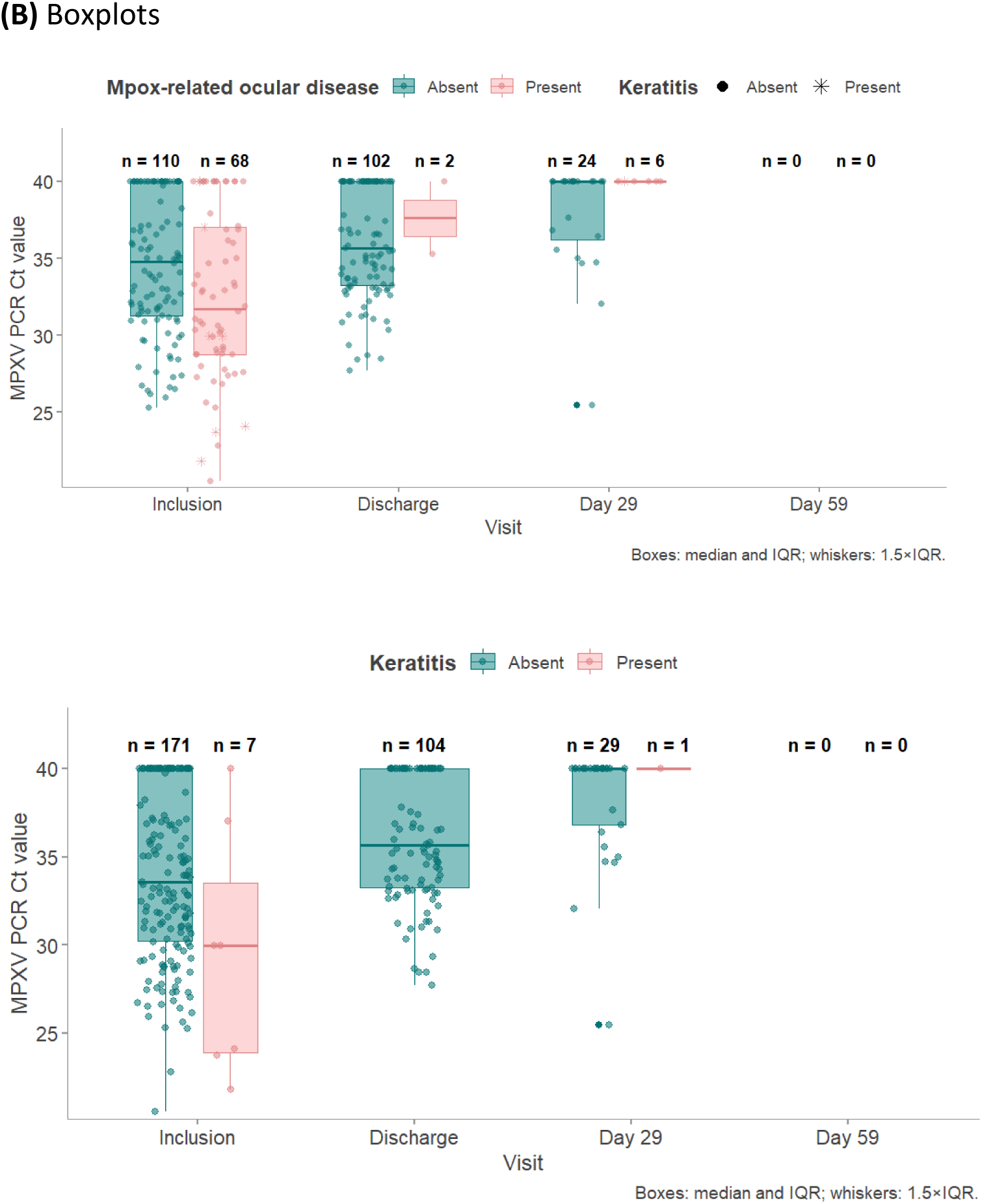
MPXV PCR Ct values over time, by mpox-related ocular disease status

In univariable regression analyses, children <5 years had a significantly higher risk of MPOXROD compared to participants >14 years (RR 2.15, 95% CI 1.48–3.13), whereas those aged 5–14 years had a comparable risk (RR 1.01, 95% CI 0.59–1.72; **Supplementary Table 3**). The presence of palpebral and periorbital mpox lesions was strongly associated with MPOXROD (RR 3.65, 95% CI 2.59–5.14, and RR 6.43, 95% CI 4.59–9.01, respectively). Severe malnutrition (RR 2.71, 95% CI 1.15–6.37, and higher viral load (lower Ct values) among PCR-positive samples (RR 1.09 per unit decrease in Ct, 95% CI 1.04–1.15) were also associated with MPOXROD. Increasing time since symptom onset was associated with a lower risk (RR 0.97 per day, 95% CI 0.96–0.98). Male sex, total number of body lesions, presence of lesions on the hands, and conjunctival PCR positivity per se were not significantly associated with MPOXROD (*p*>0.05).

In the multivariable model, only periorbital lesions (aRR 2.82, 95% CI 1.40–5.69) and severe malnutrition (aRR 5.06, 95% CI 2.25–11.38) remained independently associated with MPOXROD.

## Discussion

To our knowledge, this is the first study to prospectively conduct systematic ophthalmological assessments at multiple timepoints during an ongoing clade Ib mpox outbreak. Ocular pathology was common, affecting nearly half of individuals with MPXV infection. Conjunctivitis was the most common manifestation. However, approximately one in ten participants presented with more severe disease, including ulcerative keratitis and, more rarely, uveitis. The prevalence of specific manifestations of MPOXROD in our study exceeds those reported in a systematic review of 24 observational studies of mpox published between 1980 and 2024, which reported pooled prevalences of 8.9% (95% CI 4.4–17.1%) for conjunctivitis and 3.4% (95% CI 1.4–7.7%) for keratitis.^13^ These lower prevalence estimates likely reflect under-ascertainment in the absence of systematic dedicated ophthalmological assessment in earlier studies. Nevertheless, consistent with previous descriptions of MPOXROD, ocular involvement was largely confined to the anterior segment, particularly the ocular surface, and we did not observe rarer manifestations described in variola or vaccinia virus infections, including retinitis, chorioretinitis, optic neuropathy, or ophthalmoplegia.^13,14^

Although severe mpox-related keratitis leading to vision loss has been documented in both immunocompetent and immunocompromised individuals,^15^ the risk factors for MPOXROD remain poorly understood. In our study, children under five years had a two-fold higher risk of MPOXROD compared with older age groups, which is in line with the higher incidence of hemodynamic, pulmonary and neurological complications observed in this age group.^8^ Severe malnutrition was also independently associated with MPOXROD, suggesting that host factors influencing barrier integrity or immune responses may modify susceptibility to or severity of ocular involvement.

While the pathophysiology of MPOXROD is incompletely defined, several non-mutually exclusive mechanisms are plausible. First, ocular involvement may result from exogeneous inoculation of MPXV onto the ocular surface through infectious secretions, hands, or fomites. This mechanism is biologically plausible given the mucosal tropism of orthopoxviruses and the relative immunologic vulnerability of the conjunctiva. The higher risk of MPOXROD among individuals with periorbital lesions and the predominance of ocular surface disease supports this hypothesis, suggesting local autoinoculation or contiguous spread.

Second, MPOXROD may arise endogenously following hematogenous viral dissemination, or through secondary immune-mediated inflammation affecting ocular tissues in the context of systemic immune activation. The absence of posterior intraocular disease (e.g. retinitis, vitritis) and the low prevalence of uveitis in general argue against these mechanisms as predominant contributors to pathogenesis. The predominantly bilateral presentation noted in our study arguably could be explained by either exogenous or endogenous routes.

Once ocular involvement is established, evidence from our study and from published case reports of persistent and severe MPOXROD suggest an active role for viral replication in pathogenesis. The unique and complex immune-privilege of the ocular surface has been well-described, including in the context of viral infections.^16,17^ In our study, very low PCR Ct values were observed exclusively in individuals with active MPOXROD, particularly those with keratitis. Consistent with this, case reports, albeit in the context of clade IIb MPXV infection, have documented the persistence of MPXV DNA in ocular surface specimens for several months after disease onset, ^18–26^ with replication-competent virus cultured from corneal scrapings up to 130 days and from conjunctival swabs up to 145 days after symptom onset.^20,26^ Notably, ocular disease has been reported to develop weeks to months after resolution of cutaneous lesions,^18^ and histopathological analyses of severe mpox conjunctivitis have demonstrated necro-ulcerative inflammation with abundant intralesional orthopoxviral antigen.^27^

In contrast to most case reports, which have generally been limited to severe clinical presentations, we found that conjunctival PCR-positivity was neither sensitive nor specific for MPOXROD. Many individuals without ocular disease had detectable MPXV DNA, likely reflecting low-level, non-replicative contamination of the conjunctival sac (e.g., passive deposition of viral particles or autoinoculation). Conversely, several participants with conjunctivitis, keratitis, or uveitis had negative conjunctival PCR results. This may indicate deeper tissue involvement, immune-mediated processes, or viral replication at anatomical sites not adequately sampled by surface swabs (e.g., corneal stroma, anterior chamber). However, among PCR-positive individuals, significantly lower Ct values were strongly associated with ocular disease, consistent with a higher local viral burden, suggesting that active viral replication drives disease in at least a subset of patients with MPOXROD. Together, these findings underscore the need to interpret conjunctival PCR results in the clinical context: detection of viral DNA alone is insufficient to diagnose clinically meaningful ocular infection, whereas high viral load, when present in the context of MPOXROD, appears biologically relevant.

There are currently no antivirals proven effective for the treatment of mpox, and none have been evaluated for MPOXROD in controlled clinical trials. Tecovirimat did not shorten time to lesion resolution in two clinical trials (PALM007 and STOMP).^28,29^ Nevertheless, given their in vitro potency and in vivo activity in animal models, topical high-dose antivirals including tecovirimat, trifluridine, and (brin)cidofovir merit further evaluation.^30^ Randomized trials remain challenging: most MPOXROD is mild and self-limiting, and severe keratitis - the form most likely to benefit from antiviral therapy - is relatively uncommon. Preventive therapy is also unlikely to succeed as ocular involvement including keratitis is typically evident at first presentation. Large numbers of cases with keratitis (n>100) would be required to detect a clinically meaningful benefit in a therapeutic study, as demonstrated in trials for herpetic eye disease.^31,32^

Limitations of our study include the limited capacity to evaluate underlying immunosuppression, and the challenges of sample collection and storage in a remote setting. Temperature fluctuations during storage and transport may have contributed to variability in PCR results. Moreover, as viral culture could not be performed, we cannot determine whether detected viral DNA reflected the presence of viable virus. Our study included only patients with clade Ib MPXV infection; caution is required in generalizing our results to patients with clade Ia or clade II MPXV infections given the recognized differences in disease severity and clinical phenotypes. Key strengths include the prospective and systematic ophthalmological assessment of all confirmed mpox cases, with longitudinal clinical and virologic follow-up throughout the full clinical course, providing an unprecedented dataset to inform the prevalence, risk factors, and natural history of MPOXROD.

Ophthalmic disease in individuals affected by mpox remains a clinical challenge and appears to disproportionately affect children and individuals with severe malnutrition. Our study elucidates key aspects of the pathophysiology and natural history of MPOXROD that may inform the design of protocols for evaluating ophthalmic therapeutic approaches, including candidate topical antivirals.

## Contributors

Conceptualised the study and wrote the protocol: TK, CVD, NK, SY, VN, SSN, OTM, IC, JK, LL, and PM-K. Participant inclusion, investigation, and data collection: TK, SM, and PM. Data analysis: TK, SH, CVD, BKJ, and AT. Data management: SH, YM, and CVD. Secured funding: PM-K, LL, AWR and JK. Supervision study proceedings: SH, CVD, SSN, LL, and PM-K. Supervised laboratory procedures: TW-B, EB, SR. Writing of the initial manuscript: TK, SH, CVD, SSN, LL, and PM-K. Revised the manuscript: TK, LL, CVD, EB, TK, CVD, SM, SH, YM, PM, JCT, GM, TWB, EKL, BKJ, AT, EHV, AWR, IB, EDV, EB, SB, JCM, DBN, PK, SY, NNK, VN, OTM, IC, SSN, JK, LL, PM-K. Had full access to the data: TK, SM, YM, SH, CVD, SSN, LL, and PM-K. All authors contributed to review and approved the final version of the manuscript.

## Data sharing

De-identified participant data collected for the study will be made available from the corresponding author upon reasonable request (i.e., when ethically viable without violating the protection of participants or other valid ethical, privacy, or security concerns). For more information about the project see www.mbote-mpox.com.

## Data Availability

De-identified participant data collected for the study will be made available from the corresponding author upon reasonable request (i.e., when ethically viable without violating the protection of participants or other valid ethical, privacy, or security concerns).

## Acknowledgements

We acknowledge the dedicated staff of Kamituga General Hospital and the Alliance for International Medical Action for their continuing expert patient care. We also acknowledge the support of the Health Zone and the Provincial Health Department to facilitate setting up of the study and the Kamituga General Hospital for generously disposing the study team of necessary infrastructure to pursue study activities. Finally, we thank our colleagues from ITM Kinshasa office for administrative and logistical support.

## Use of Artificial Intelligence

During the preparation of this work, the authors used ChatGPT-5.2 as a language editing tool. After using this tool, the authors reviewed and edited the content as needed and take full responsibility for the content of the publication.

