## Supplement for "Prevalence, Characteristics and Evolution of Mpox-Related Ophthalmic Disease: A Prospective Cohort Study in South-Kivu, Democratic Republic of the Congo (MBOTE-EYE)"

**Supplementary Table 1: Prevalence of eye disease, per participant, by age category.**

| | Age < 5 years | Age 5 – 14 years | Age > 14 years |
|---|---|---|---|
| **Affection of periorbita & palpebra, n/N (%; 95% CI)** | | | |
| Periorbital mpox lesions | 35/105 (33.3) | 14/56 (25.0) | 30/149 (20.1) |
| Blepharitis | 12/105 (11.4) | 4/56 (7.1) | 8/149 (5.4) |
| Palpebral mpox lesions | 31/105 (29.5) | 10/56 (17.9) | 25/149 (16.8) |
| **MPOXROD (mpox-related ocular disease), n/N (%)** | 65/105 (61.9) | 20/56 (35.7) | 49/149 (32.9) |
| Conjunctivitis | 62/105 (59.0) | 20/56 (35.7) | 50/149 (33.6) |
| Without conjunctival mpox lesion(s) | 59/62 (95.2) | 17/20 (85.0) | 43/50 (86.0) |
| With conjunctival mpox lesion(s) | 3/62 (4.8) | 3/20 (15.0) | 7/50 (14.0) |
| Scleritis | 0/105 (0.0) | 0/56 (0.0) | 0/149 (0.0) |
| Keratitis | 16/105 (15.2) | 3/56 (5.4) | 10/149 (6.7) |
| Non ulcerative | 2/16 (12.5) | 0/3 (0.0) | 0/10 (0.0) |
| Ulcerative | 14/16 (87.5) | 3/3 (100.0) | 10/10 (100.0) |
| Anterior uveitis | 0/105 (0.0) | 0/56 (0.0) | 2/149 (1.3) |
| Intermediate/posterior uveitis | 0/105 (0.0) | 0/56 (0.0) | 0/149 (0) |
| Affection of the optic nerve | 0/105 (0.0) | 0/56 (0.0) | 0/147 (0)[$] |
| **Functional outcome at day 59, n/N (%)** | | | |
| Snellen score < 8/10 | 13/105 (12.4) | 14/56 (25.0) | 23/149 (15.4) |
| Snellen score < 3/10 | 0/105 (0.0) | 2/56 (3.6) | 5/149 (3.4) |

[$] Excluding two participants with a prior history of glaucoma and pallor of the optic nerve at enrolment

**Supplementary Table 2: Result of MPXV-PCR on conjunctival swabs, by eye, at presentation and during follow-up**

| | Inclusion | Discharge | Day 29 | Day 59 |
|---|---|---|---|---|
| **Number of available swabs** | 178 | 106 | 30 | 0 |
| **Number of valid test results** | 178 | 104 | 30 | 0 |
| **Number of participants with valid test results** | 89 | 52 | 15 | 0 |
| **Proportion testing positive, n/N (%)** | 132/178 (74.2) | 65/104 (62.5) | 9/30 (30.0) | NA |
| **Median ROX Ct value (IQR)** | 33.4 (29.7 – 40.0) | 35.7 (33.2 – 40.0) | 40.0 (37.0 – 40.0) | NA |
| **Median ROX Ct value (IQR) among PCR-positive samples** | 31.7 (28.9 – 34.7) | 33.8 (32.7 – 35.3) | 35.0 (34.7 – 36.4) | NA |

Conjunctival swabs were collected from 29/10/2024 until 5/12/2024

**Supplementary Table 3: Factors associated with MPOXROD**

| | Univariable model | | Multivariable model* | |
|---|---|---|---|---|
| | **RR (95% CI)** | ***p*-value** | **aRR (95% CI)** | ***p*-value** |
| Age > 14 years | *Reference category* | - | - | - |
| Age 5-14 years | 1.005 (0.586–1.723) | 0.987 | 1.204 (0.514–2.819) | 0.669 |
| Age < 5 years | **2.154 (1.482–3.131)** | **0.000058** | 1.889 (0.843–4.234) | 0.122 |
| Sex: female | *Reference category* | - | - | - |
| Sex: male | 1.216 (0.857–1.723) | 0.273 | - | - |
| Presence of palpebral mpox lesions | **3.649 (2.592–5.136)** | **<0.00001** | 1.153 (0.644–2.065) | 0.633 |
| Presence of periorbital mpox lesions | **6.428 (4.587–9.007)** | **<0.00001** | **2.821 (1.398–5.692)** | **0.00378** |
| Presence of mpox lesions on the hands | 0.936 (0.628–1.395) | 0.745 | - | - |
| Malnutrition[£]: no/moderate | *Reference category* | - | - | - |
| Malnutrition[£]: severe | **2.709 (1.153–6.366)** | **0.0222** | **5.064 (2.253–11.382)** | **0.000087** |
| Lesion severity: mild | *Reference category* | - | - | - |
| Lesion severity: moderate | 0.875 (0.521–1.469) | 0.614 | - | - |
| Lesion severity: severe | 1.144 (0.685–1.91) | 0.607 | - | - |
| Lesion severity: grave | 1.351 (0.795–2.296) | 0.266 | - | - |
| Conjunctival swab PCR negative | *Reference category* | - | - | - |
| Conjunctival swab PCR positive | 1.417 (0.887–2.264) | 0.144 | 0.603 (0.3–1.208) | 0.154 |
| Inverted Ct-value of PCR-positive swabs ** | **1.089 (1.036–1.146)** | **0.000843** | 1.061 (0.991–1.135) | 0.0908 |
| Time since onset of symptoms (days) | **0.966 (0.955–0.977)** | **<0.00001** | 0.96 (0.91–1.013) | 0.134 |

* The multivariable model included all variables with *p*-value < 0.2 in univariable analysis, including 312 observations from 88 individuals. Associations with *p*-value < 0.05 are displayed in bold.

** represented by the interaction term between inverted Ct-values and PCR positivity

[£] Moderate malnutrition was defined as MUAC 11.5–12.5 cm in children <5 years; BMI-for-age z-score −3 to −2 SD in participants aged 5–19 years; and BMI 16–18 $kg/m^2$ in adults >19 years. Severe malnutrition was defined as MUAC <11.5 cm in children <5 years; BMI-for-age z-score <−3 SD in participants aged 5–19 years; and BMI <16 $kg/m^2$ in adults >19 years.
